# COVID-19 Infection Among Healthcare Workers in a National Healthcare System: the Qatar Experience

**DOI:** 10.1101/2020.08.09.20170803

**Authors:** Jameela Alajmi, Andrew M. Jeremijenko, Joji C. Abraham, Moza Ishaq, Elli G. Concepcion, Adeel A. Butt, Abdul-Badi Abou-Samra

## Abstract

The study was conducted at Hamad Medical Corporation in Qatar, a national healthcare system with 14 hospitals and over 28,000 employees. A total of 16,912 staff members were tested for SARS-CoV-2 between March 10 and June 24, 2020 with 1,799 (10.6%) testing positive. Nurses and midwives had the highest number of infections (33.2% of all infected HCWs) followed by non-clinical support service staff (31.3%), administrative staff (14.6%), allied health professionals (12.7%), physicians (5.2% and other clinical support staff (2.9%). Among 671 infected HCW surveyed by the infection prevention and control team immediately after the positive COVID-19 test was reported, exposure to a family member or roommate with confirmed infection each were reported by 9.5%. Two-thirds of the infected HCWs were symptomatic with fever (34.6%), cough (32.2%) and sore throat (15.8%) being the most commonly reported symptoms. Among the survey respondents, 78 (11.6%) were hospitalized, 9 (1.3%) required supplemental oxygen, 4 (0.6%) were admitted to the intensive care unit) and 2 (0.3%) required mechanical ventilation. There were no deaths. To understand the transmission dynamics and impact of facility designation as COVID-19 or non-COVID-19 facility, we conducted a focused follow-up telephone survey on 393 COVID-19 positive HCW 1-6 weeks after diagnosis. Only 5% of respondents reported acquiring the virus from working at a COVID-19 designated facility while the remaining 95% reported working at a non-COVID-19 facility and acquired the infection from accidental exposure to a colleague (45%) or to a patient (29%). Among infected HCW at COVID-19 designated facilities, 82% reported used full PPE at all times while 68% of infected HCW at non-COVID-19 facilities reported using PPE as directed.

## Introduction

Since the identification of first case cluster in Wuhan, China in December 2019, the COVID-19 pandemic has swept the entire world. The pandemic has overwhelmed hospital capacity and existing healthcare resources in many countries. Healthcare workers (HCW) are a particularly high risk group due to their close interactions with infected persons as well as lack or deficiency of personal protective equipment in many settings. The rate of infection in HCW is reported to vary between 3-17% and varies according to the history and degree of exposure and presence of symptoms.^1-4^ It is important to understand the prevalence and risk factors for COVID-19 infection among HCW due to the potential to transmit infection to vulnerable patients, and since a further depletion of the workforce due to infection among the HCW can lead to critical shortages and adversely impact patient care. Our aim was to determine the prevalence of COVID-19 infection in HCW in a national healthcare system and to understand the risk factors for infection.

## Methods

The study was conducted at Hamad Medical Corporation in Qatar between March 10, 2020 and June 24, 2020. Hamad Medical Corporation is the public healthcare delivery organization providing approximately 85% of the acute care inpatient bed capacity in the State of Qatar. There are 14 hospitals in the organization with a total employed staff of more than 28,000 persons. All hospitals are accredited by the Joint Commission International and the central lab is accredited by the College of American Pathologists. There is a single electronic health record system (Cerner, Kansas City, USA) across all facilities and patients and staff retain the same unique hospital identification number across the system.

The first case of COVID-19 in Qatar was identified on February 28, 2020 in a returning traveler. However, screening of potential high risk and symptomatic persons had begun in early February. Testing capacity in Qatar was ramped up rapidly and currently Qatar ranks among the countries with highest per capita testing in the world. Screening of HCW began in mid-February 2020, with initial screening performed on symptomatic HCW and those with a history of any contact with a confirmed or suspected case. Testing for COVID-19 was performed using a deep nasopharyngeal and a concomitant throat swab by trained professionals. Validated RT-PCR was performed at a single national reference lab to confirm infection. Beginning in April 2020, all HCW in intensive care units and emergency departments were required to undergo point-of-care testing using commercial antibody kits (company name) every two weeks. Those testing positive for IgM provided a nasopharyngeal and throat swab for RT PCR testing.

All HCW at any of the HMC facilities with a RT PCR positive for COVID-19 on a nasopharyngeal swab were eligible to be included in this study. We retrieved demographic and employment related information from the electronic health records and employment records in the Human Resources department. Trained infection control practitioners at each facility approached all positive HCWs through email and follow-up telephone calls to gather data regarding exposure history, use of personal protective equipment and presence of symptoms. The WHO COVID-19 staff exposure risk assessment tool was used as a template to gather data. Hospitalization data, including admitting unit and need for supplemental oxygen were extracted from the electronic health records.

The study was approved by the Institutional Review Board of Hamad Medical Corporation with a waiver of informed consent under a pandemic response framework adopted by the institution.

## Results

A total of 16,912 staff members were tested for SARS-CoV-2 between March 10 and June 24, 2020 with 1,799 (10.6%) testing positive. The median age (IQR) was 39 (33,48) years and 65.6% were male. The most common nationalities affected were Indian (50.8%), Filipino (14.0%), Qatari 6.1% and Egyptian (5.6%), reflecting the overall demographic profile of the HMC employees. At least one national from 27 additional countries were infected collectively amounting to 8.2% of the infected HCWs. **(Table 1)** Nurses and midwives had the highest number of infections (33.2% of all infected HCWs) followed by non-clinical support service staff (31.3%), administrative staff (14.6%), allied health professionals (12.7%), physicians (5.2% and other clinical support staff (2.9%).

**Table 1.**
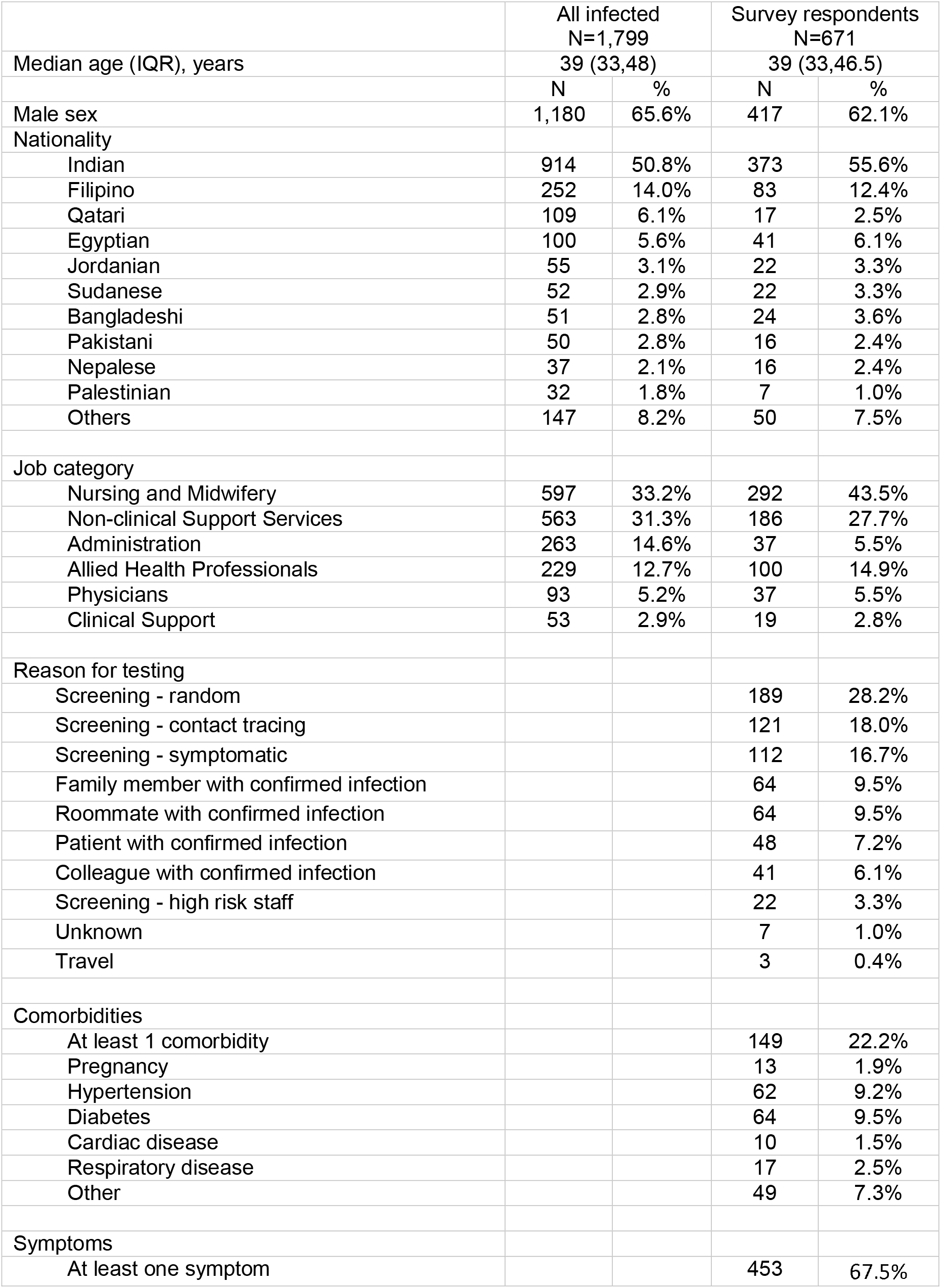
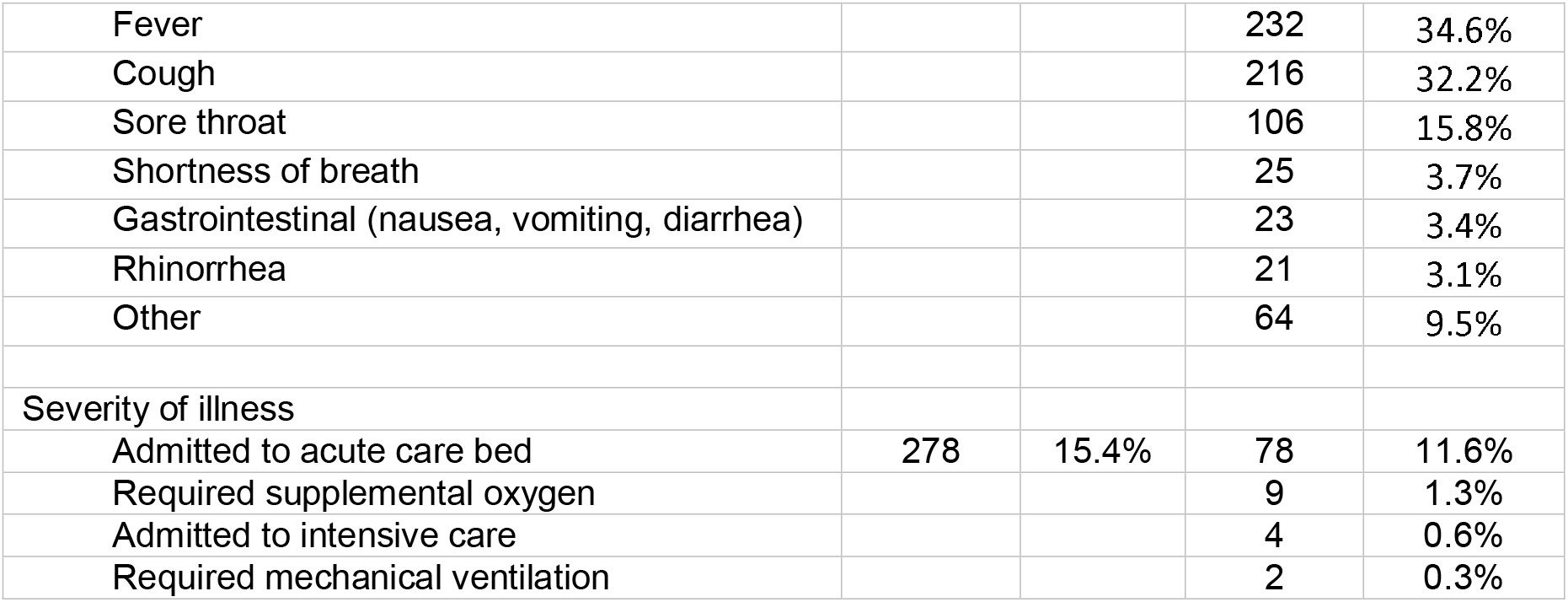
Baseline characteristics.

A total of 671 infected HCW were surveyed by the infection prevention and control team immediately after the positive COVID-19 test was reported. Demographics of this groups were similar to the overall group. The most common reasons for testing were random screening (28.2%), screening as a part of contact tracing (18.0%) or presence of symptoms suggestive of COVID-19 infection (16.7%). Exposure to a family member or roommate with confirmed infection each were reported by 9.5%. Two-thirds of the infected HCWs were symptomatic with fever (34.6%), cough (32.2%) and sore throat (15.8%) being the most commonly reported symptoms. Among all infected HCWs, 278 (15.4%) were hospitalized. Among the survey respondents, 78 (11.6%) were hospitalized, 9 (1.3%) required supplemental oxygen, 4 (0.6%) were admitted to the intensive care unit) and 2 (0.3%) required mechanical ventilation. There were no deaths.

Early in the pandemic preparedness phase, Qatar had designated certain hospital and healthcare facilities to be non-COVID-19 facilities, with the intention of cohorting COVID-19 patients at specific designated facilities for efficiency, optimal utilization of resources, and improved infection prevention. To understand the transmission dynamics and impact of facility designation as COVID-19 or non-COVID-19 facility, we conducted a focused follow-up telephone survey on 393 COVID-19 positive HCW 1-6 weeks after diagnosis. Only 5% of respondents reported acquiring the virus from working at a COVID-19 designated facility while the remaining 95% reported working at a non-COVID-19 facility and acquired the infection from accidental exposure to a colleague (45%) or to a patient (29%). Among infected HCW at COVID-19 designated facilities, 82% reported used full PPE at all times while 68% of infected HCW at non-COVID-19 facilities reported using PPE as directed.

## Discussion

Protection of HCW from infection is critical for resilience of the health system facing a major pandemic such as COVID-19. However, despite all efforts to protect HCW, some exposure is inevitable. Such exposure can occur at the workplace or outside the work environment in the community.

To reduce workplace exposure, increase efficiency, and pooling of resources, Qatar decided to designate certain facilities to be non-COVID-19 facilities before the pandemic hit the country. It was therefore anticipated that HCW working at non-COVID-19 facilities would be at a very low risk for infection acquired at the workplace. It was therefore surprising that 95% of the infected HCW were assigned to a non-COVID-19 facility and 72% of these reported exposure to a coworker or patient as the source of infection. Possible reasons for this include a lower rate of adherence to prescribed PPE at non-COVID-19 facilities, complacency with strict infection prevention precautions, and unrecognized infection among patients and coworkers. Conversely, at COVID-19 designated facilities, HCWs may have been more adherent to PPE use and infection prevention measures since most, if not all patients in these facilities were known to be COVID-19 infected.

Our findings underscore the need for strict PPE use and infection control measures regardless of workplace environment and whether any patients or coworkers are known to be COVID-19 infected. This is critical to reduce infection rates among HCWs and to ensure reliable availability of the workforce in times of critical need like the current COVID-19 pandemic.

## Data Availability

Data not publicly available.

## Disclaimer

The views presented in this manuscript are those of the authors and do not necessarily represent the views or official policy of Hamad Medical Corporation or the Ministry of Public Health, Qatar.

## Author contributions

Study design: JA; AAB; AA;

Data acquisition: JA; AA; JCA

Data analysis: AAB; AA;

Manuscript writing: AAB; AA;

Critical review and major scientific input: JA; AMJ; JCA; MI; EGC; AAB; AA

